# Development and implementation of a nowcasting method for the syndromic surveillance of severe acute respiratory infections (SARI) in Germany

**DOI:** 10.1101/2025.10.28.25338979

**Authors:** Felix Günther, Kristin Tolksdorf, Ulrich Reinacher, Ekkehard Schuler, Silke Buda, Frank Sandmann

## Abstract

**Background:** In Germany, diagnosis-based hospital surveillance of severe acute respiratory infections (SARI) incidence is operated weekly. However, diagnosis data from the most recent weeks is provisional and may be subject to systematic changes that we wish to account for to achieve a timely assessment of SARI trends.

**Methods:** We developed a novel nowcasting method based on (i) the already available data for the current week and (ii) historic changes in the data in weeks following initial reporting. The available weekly data were rescaled using multiplicative factors based on the historic patterns, to obtain the expected final incidence. We fit a parametric Beta distribution for observed upward changes in reported incidence, and a mixture distribution in case of the existence of downward changes, e.g., due to later corrections in diagnoses. We evaluated the model performance for the respiratory winter season 2024/2025.

**Results:** The average weekly SARI incidence upon initial estimation in the first week after hospitalisation was 79.4% (median) of the incidence five weeks after hospitalisation, which was considered final. It increased to 95.1% in the second week after hospitalisation. The nowcast predictions matched the final incidences well and were able to indicate trends in real-time. Prediction intervals were well-calibrated and nowcasting can be performed in subgroups given sufficient case numbers.

**Conclusions:** The SARI surveillance in Germany achieved a high completeness already in the initial weeks after hospitalisation. The newly-developed nowcasting method yielded robust results with reliable uncertainty quantification. In combination, this facilitates a close to real-time assessment of SARI hospitalisation incidence in Germany.

## Introduction

The Robert Koch Institute (RKI) is tasked with the real-time surveillance of acute respiratory infections (ARI) activity and severity in Germany to provide situational awareness and policy advice during seasonal epidemics and pandemics caused by respiratory pathogens. To this end, multiple surveillance systems have been implemented and operated at RKI based on statutory case reporting of respiratory pathogens like Influenza virus, SARS-CoV-2 and RSV, virological sentinel surveillance data, wastewater surveillance data, and syndromic sentinel surveillance data. The results of these surveillance systems are assessed and summarised in national situation reports^1^ and reported to supranational health authorities like ECDC^2^ and WHO^3^ year-round. One of the key sentinel systems is the ICD-10 code-based hospital surveillance of severe acute respiratory infections (ICOSARI)^4^. The SARI surveillance is a hospital-based sentinel system that complements the syndromic ARI surveillance systems on population level and in primary care with a representative estimate of the weekly incidence of severe ARI (SARI) requiring hospital admission in Germany.

The SARI surveillance uses electronic health record data, specifically main discharge diagnosis codes and also preliminary diagnosis codes, which enable a (very) timely monitoring based on an initial reporting of a weekly hospitalisation incidence already in the first week after hospitalisation. However, preliminary diagnosis data can change during the duration of hospital stay and more diagnosis codes can be added. Typically, such retrospective changes of the initial reported data result in an increase in SARI cases due to added diagnosis codes (“right-truncation”). However, also decreases in SARI case numbers are possible. These changes and updates of data can complicate assessment and identification of recent trends in real-time surveillance.

Delays and retrospective updates occurring at various stages of data gathering and reporting are well known and common to many sources of incidence data in infectious diseases surveillance. In response, statistical approaches have been developed to adjust for such delays. These approaches are referred to as nowcasting methods and have often been developed in the context of specific data streams and for different pathogens, for example in the surveillance of measles, influenza like illness, dengue, Shiga toxin-producing *Escherichia coli*, MPox, and were also of particular relevance for various indicators during the SARS-CoV-2 pandemic^5-12^.

In this work, we aimed to systematically investigate the magnitude of right-truncation in the SARI hospitalisation incidence and develop a robust nowcasting method to adjust for the truncation and the changes of the incidence data following their initial reporting. We evaluated the performance of the model during the 2024/2025 ARI winter season and investigated the benefits of applying the methods for real-time monitoring.

## Methods

### Data

In 2015, the RKI started to collaborate with a private hospital network to develop a SARI sentinel surveillance system using case-based data on ICD-10 codes^4^. Since then, anonymized patient data originating from quality assurance reports from a network of private hospitals belonging to the *HELIOS Kliniken GmbH* are sent to the RKI on a regular basis. At RKI, these data are processed to estimate a weekly SARI incidence by comparing incident (i.e., newly admitted) patients with ICD-10 codes J09-22 in their main diagnosis to the catchment population of the sentinel hospitals^13^. Processing of case-based data facilitates also stratified incidence estimation, e.g. for certain age groups. Initially, discharge diagnosis codes were used for individuals with a hospital stay of less than one week, and incidence was reported in the second week after hospitalisation. Since then, improvements have been made to data transmission and processing, so that data is now transmitted daily rather than weekly and preliminary main diagnosis codes are used, as long as no discharge data is avaible. Thus, the SARI case definition had been changed to include not only patients with a main discharge diagnosis of J09-22, but also patients who are still in hospital and have an according preliminary main diagnosis of J09-22, without any further restriction regarding duration of hospitalisation.

The weekly SARI incidence estimate from the hospital sentinel is published year-round as part of weekly national situation reports every Wednesday (“ARI weekly reports”^1^), visualised in a dashboard^14^published in a public repository^15^ and also uploaded to the European Surveillance System (TESSy), a central data platform hosted by ECDC^16^, from where it is included in the European Respiratory Virus Surveillance Summary (ERVISS)^2^. For all of these reports, the reported estimates are based on the data sent from hospitals to a datacentre on Monday night, that are forwarded to RKI on Tuesday morning where they are processed during the day. Weekly SARI incidences are calculated by using the number of SARI cases, that were admitted to hospital in a given week, and the catchment population of the sentinel hospitals. The catchment population of the sentinel is estimated by applying the proportional flow method, where the number of all admitted patients in the sentinel per year, age group and region is set into relation to the number of admitted patients throughout Germany^13^.

The changes made to the data transfer and processing increased timeliness of the available data substantially. SARI cases could now be initially reported in the first week after hospitalisation. Nevertheless, the data and estimated incidence for a given week may still be revised in the first few weeks after the initial estimation and reporting. Usually, the estimates for the SARI incidence of a certain week increase after initial reporting as the data transfer for additional cases diagnosed with ICD-10 codes for SARI occurs over the following days.

### Changes in incidence after initial reporting (completeness)

We conducted a descriptive analysis of changes in the weekly incidence after initial reporting in the first week after hospitalisation during the 2024/2025 winter season (defined as running from calendar week 40 in 2024 to calendar week 20 in 2025; in short: 2024-W40 – 2025-W20). After an initial evaluation of such changes, we defined the final incidence for each week as the estimated incidence for the respective week four weeks later (i.e., five weeks after hospital admission), as no systematic increase or decrease in the incidence estimate was observed thereafter. To describe changes in incidence over time, we then compared the incidence estimated in the first week after hospital admission (i.e. at the time of initial estimation and reporting) with the final incidence (five weeks after hospital admission) based on their relative proportions for each week of the 2024/2025 winter season. Expressed as a percentage, this proportion can be interpreted as the *completeness* of the incidence upon initial estimation and reporting. Following the same procedure, we also investigated the completeness of the incidence during the weeks following initial reporting (e.g., comparing the estimated incidence in the second, third week, or fourth week after hospitalisation to the final incidence). As the incidence estimate for a specific week is based not only on final discharge diagnosis codes, but also on preliminary diagnosis codes of still hospitalised patients that may be revised, such revisions in diagnoses may also lead to downward corrections in the estimated final incidence and the *completeness* of the incidence estimate for certain combinations of hospitalisation and reporting week could be larger than 100% (of the final incidence for the respective week of hospitalisation).

We analysed the changes in completeness of SARI incidence for Germany overall as well as in six different age groups (age 0-4, 5-14, 15-34, 35-59, 60-79, and 80+ years) to investigate potential differences between the age groups.

### Developed nowcasting method

We developed a nowcasting method to estimate the final SARI incidence for the most recent and the three previous weeks based on the already-available incidence data and historic information about changes after initial reporting. Conceptually, the idea is to investigate the (multiplicative) changes of the SARI incidence in the initial reporting week (first week after hospitalisation), or two or more weeks after hospitalisation compared to the final incidence (defined in our cases as the incidence D=5 weeks after hospitalisation). The currently available incidence data is then rescaled according to the historic changes to obtain a “nowcasted” estimate of the expected *final* incidence for all weeks up to the most recent week.

With more technical detail, based on data available in a certain week *T*, our developed nowcasting method works as follows: In a first step, we define a maximum delay of D weeks after which no further changes in the incidence are assumed. The reporting pattern can then be captured in a 2-dimensional array: Let *I*_*t,d*_ be the incidence of week *t*= 1, …, *T* − 1, reported *d*= 1, …, *D*, …, *T* − 1 weeks after hospitalisation in week *t*. Note that *t* is a consecutive number (integer) and is not corresponding to the numeric value of the calendar weeks. If we want to perform nowcasting, e.g., for calendar week 50 in a certain year, based on data of *T*= 30 weeks, the week *t*= 1, corresponds to calendar week 21 of the same year and *I*_1,5_ is the incidence reported for calendar week 21 based on data available in calendar week 26 (i.e., five weeks later).

In a specific week *T* (i.e., the most recent, “current” week) the incidence data *I*_*t,d*_ is partly unavailable, the most recent incidence data available for week *t*= 1, …, *T* − 1 (i.e., the current incidence time series) corresponds to *I*_*t,T*−*t*_. For all weeks *t* ≤ *T* − *D*, the incidence considered as final (*I*_*t,D*_) is already available, for all weeks *t* > T − *D*, we expect further changes.

In a second step, we calculate *J* empirical multiplicative changes comparing the incidence *d*= 1, …, *D* − 1 weeks after hospitalisation to the final incidence *D* weeks after hospitalisation based on the most recent *J* weeks for which the final incidence is already available. These multiplicative factors are therefore defined as *κ*_*t,d*=_ *I*_*t,d*_/*I*_*t,D*_, for *t*= *T* − *D* − *J* + 1, …, *T* − *D* and *d*= 1, …, *D* − 1. All required incidence data can, e.g., be extracted from snapshots of weekly incidence time series as reported in consecutive weeks.

In a third step, we fit *D* − 1 independent parametric distributions 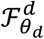 to the *J* multiplicative changes *κ*_*t,d*_ that are supposed to characterize subsequent changes of incidence data *d*= 1, …, *D* − 1 weeks after the week of hospitalisation, where *θ*_*d*_ is a parameter vector of the distribution. In the simplest situation, all *κ*_*t,d*_ for a certain *d* lie between zero and one (i.e., the reported incidence for week *t* at week *t* + *d* is a fraction of the final incidence of week *t* known at week *t* + *D* smaller than one). In this situation, we fit a parametric Beta distribution to the observed values of *κ*_*t,d*_. In practice, some of the values of *κ*_*t,d*_ can already equal one (i.e., complete reporting of the incidence for week *t* already *d* < *D* weeks after initial reporting) or they can be even bigger than one (in case of subsequent downward-corrections of the dataset when preliminary diagnostic codes were revised at a later point in time). For such situations, we fit a more complex mixture distribution 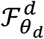 to the observed multiplicative changes (see Supplementary Text 1).

In a fourth step, we draw *L* (e.g., *L*= 1000) independent random multiplicative factors 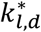 from each of the *D* − 1 fitted mixture distributions. To account for the estimation uncertainty in the parameters of the mixture distribution, we first draw a random realization of the parameter vector 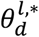 with expectation 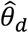 and covariance derived from the likelihood-based fitting of the parametric (mixture) distribution, and then sample a random multiplicative factor 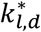 from the distribution 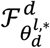, with *l*= 1, . . ., *L*.

To obtain the nowcasted incidence time-series, we construct L incidence trajectories as

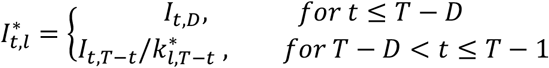

The nowcasted time-series Î_*t*_ can then be derived from the weekly point-wise mean of the trajectories/samples 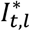, *t*= 1, …, *T* − 1. Quantiles of the predictive distribution correspond to the empirical quantiles of the samples and are used for constructing prediction intervals.

Results for age groups were estimated independently using the same approach. Results of the nowcast were provided weekly internally during the study period to improve the epidemiological SARI assessment in the national weekly ARI report.

### Illustration of results from the nowcasting method

In this paper, we illustrate the nowcasting method by applying it to the SARI incidence data that was available at different time points during the 2024-2025 ARI season. As a main example, we used calendar week 8, 2025, when SARI incidence peaked during the seasonal influenza wave. We show results also based on five additional data snapshots to illustrate the additional information provided by nowcasting when incidence was increasing or decreasing to examine the extent to which the preliminary and nowcasted incidence inform real-time trend assessment. We performed the nowcast for the overall SARI incidence, as well as separately in six age groups.

### Performance evaluation

The developed nowcasting method was prospectively piloted internally at RKI from week 3 in 2025 in real-time, with a weekly interpretation of results and monitoring of the model performance. To assess the performance of the nowcasting method more robustly, we also performed a retrospective evaluation over the whole respiratory infection season 2024/2025. For this, we performed the nowcasting for each week from calendar week 40, 2024 up until calendar week 20, 2025 and compared the predictions to the final incidence data. We evaluated the performance in terms of the mean absolute error (MAE), root mean squared error (RMSE), and the scaled mean absolute percentage error (sMAPE) of the nowcast point predictions. To investigate uncertainty quantification, we also evaluated empirical coverage frequencies of prediction intervals, as well as the weighted interval score (WIS). Evaluation was performed separated by nowcast horizon, e.g., the nowcasted incidence in the week of initial reporting, one week after initial reporting, and so on. We evaluated the performance for the overall incidence, as well as the incidence in age groups. For the retrospective evaluation, the calendar weeks 51 and 52 in 2024 were excluded as no data was processed and reported in these weeks, and hence no SARI incidence could be robustly estimated during the Christmas and New Year holidays.

All analyses were performed in R based on a custom implementation of the nowcasting method using functionalities of the gamlss package^17^ for estimating distributional parameters. The analyses are implemented in a targets pipeline^18^ and code is published alongside this article in a public code repository.

## Results

### Changes in incidence in the weeks following initial reporting

During the 2024/2025 winter season, the SARI incidence estimates generally increased in the weeks following initial reporting. Figure 1A exemplifies this for the incidence data up to calendar week 8, 2025 (February 17-23, 2025). Based on the incidence data reported on Wednesday, February 26 (first reporting of incidence up to and including calendar week 8), there was a stabilization and even a small decrease in the SARI incidence since week 6, 2025 (grey bars in Fig. 2A). However, the final data after 4 additional weeks showed a slightly different pattern: although the increase in incidence from calendar weeks 4 and 5 had slowed, there had been at most a stabilization of incidence by calendar week 8, but not yet a decline (Fig. 2A, orange line). Compared to the final data, only 79.4% of the SARI incidence was reported in the initial reporting week for calendar week 8. For calendar week 7, this proportion was already 95.5% (first week after initial reporting) and 98.1% for calendar week 6 (second week after initial reporting).

**Figure 1.**
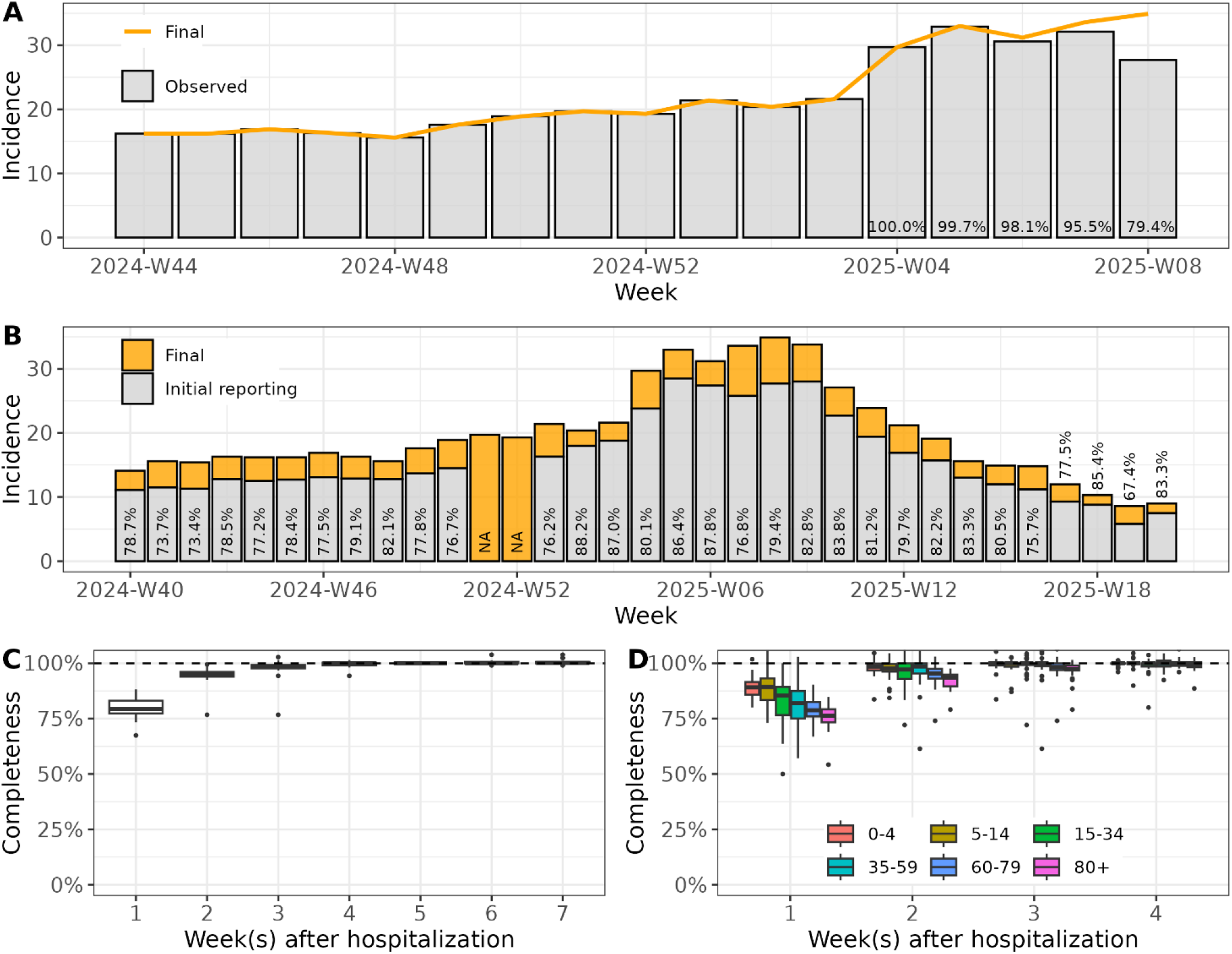
Descriptive visualization of changes in the SARI incidence data following initial reporting. Panel A shows the SARI incidence data up until calendar week 8, 2025 (February 17-23, 2025) as available on February 25, 2025 (calendar week 9, grey bars) as well as the “final” incidence time series available 4 weeks later (in calendar week 13, orange line), numbers in bars correspond to the fraction of the initial data compared to the final incidence (“completeness”, in percent). Panel B shows the reported incidence upon initial reporting as well as the final incidence for the whole winter season 2024/2025; numbers in bars correspond to the fraction of the initial data compared to the final incidence (“completeness”, in percent). Panel C shows the reported incidence completeness of reported incidence upon initial reporting (in the first week after hospitalisation) and in the following weeks for the whole winter season 2024/2025 (calendar week 40, 2024 – calendar week 20, 2025). Panel D shows this completeness for six separate age groups.

**Figure 2.**
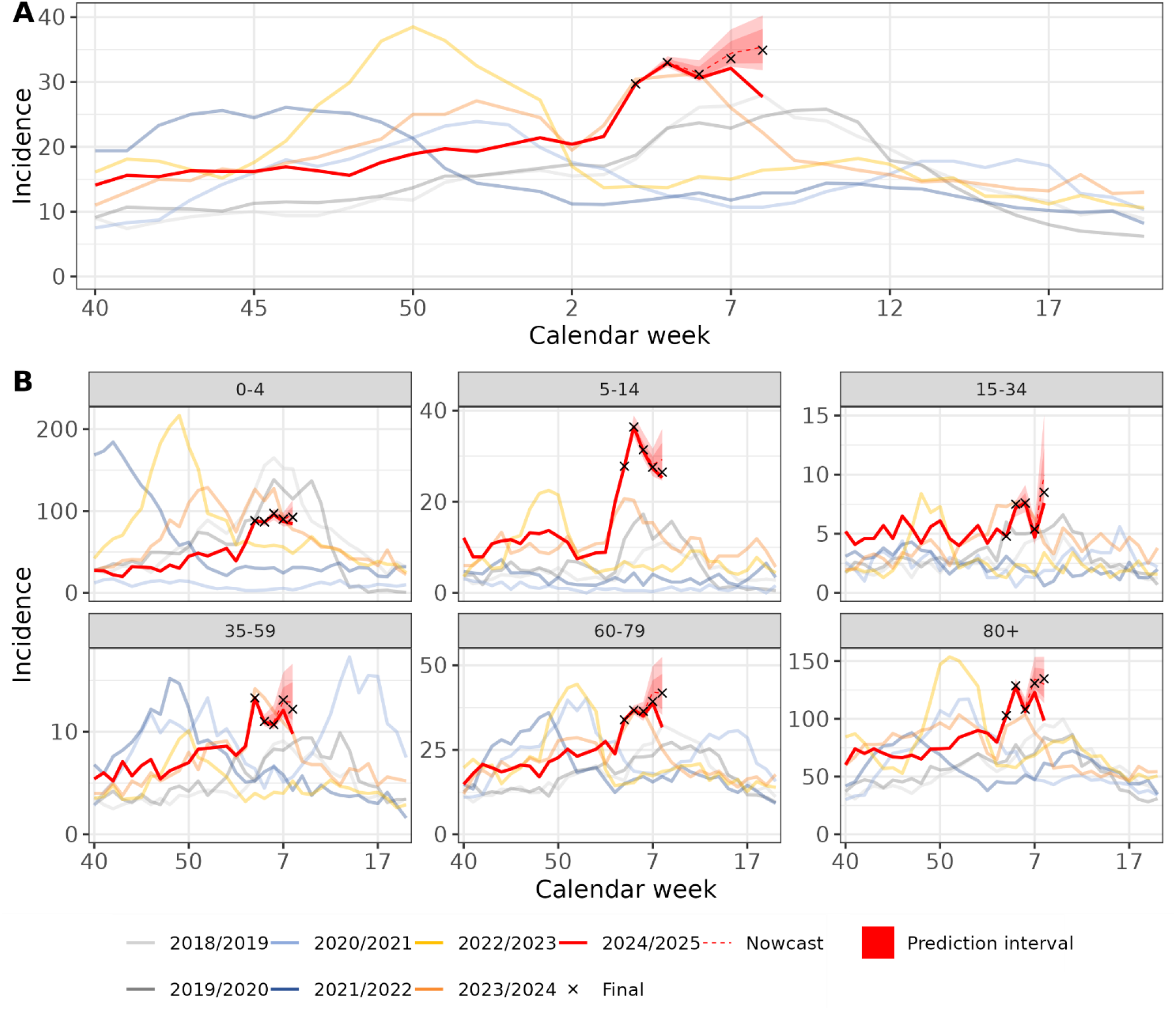
Illustration of nowcast method for the SARI incidence up to calendar week 8, 2025 (total; and by age). Panel A shows the reported incidence data by calendar weeks in years (color-coded) as well as the nowcast estimate of the expected final incidence (red dotted lines) and corresponding 80% and 95% prediction intervals (ribbons). Black crosses show the final incidence time series that is unavailable at the time of nowcasting. Panel B shows results in age groups.

The proportion of the final incidence initially reported was relatively stable during the weeks of the 2024/2025 winter season, ranging from 67.4% to 88.3% in the calendar weeks 2024-W40 to 2025-W20, with the exception of the two calendar weeks 2024-W51 and W52 around Christmas and New Year in which no incidence was reported. The median incidence reported at the time of initial reporting was 79.4% of the final incidence (25%/75% quantile: 77.3%, 83.1%; Fig 1B and C).

In the first week after initial reporting, the median reported incidence was 95.1% (25%/75% quantile: 94%, 96.1%) compared to the final incidence. This proportion continued to increase, and 4 weeks after initial reporting and beyond, there were generally no substantial changes in the estimated SARI incidence anymore (Figure 1C). In nine of the 31 weeks included (29%), the incidence data two or three weeks after initial reporting was slightly higher than the “final” incidence four weeks after initial reporting (maximum of 3% for reference week 2025-W03: incidence of 22.2 two weeks after initial reporting, final incidence of 21.6).

Looking at the incidence data by age group, reporting patterns were in general similar, although the incidence data in the younger age groups tended to be more complete at initial reporting compared to the older age groups with the median completeness ranging from 89.1% to 76.3% (Fig. 1D). Moreover, variability in the completeness over the weeks was larger within the age groups, as population size decreased with stratification and retrospective changes in single cases had a larger impact, especially when the SARI incidence was rather low. Thus, also downward changes in the estimated SARI incidence occurred more frequently, e.g., for 16 out of 31 weeks in the age groups 35-59 and 60-79 years.

### Results of the nowcast at different time points of the 2024/2025 ARE season

When applying the nowcasting method to the initially reported incidence data for calendar week 8, 2025, we observed that the nowcast predictions captured the trends in the final incidence time-series well, and was therefore able to adjust for changes in the incidence time series after initial reporting (Fig. 2A). Although the nowcast was subject to relevant uncertainty (especially for the last calendar week 8), it already indicated that the incidence was still stable or slightly rising up to calendar week 8 and that the peak of the SARI winter wave had not yet been passed at that point in time.

We also performed nowcasting separately in six age groups. Despite low incidence in some of the age groups (e.g., 15-34 and 35-59) the nowcast performed well with trends in final incidence being captured adequately. Relative uncertainty in the nowcast prediction tended to increase with lower incidence. The 95% and 80% prediction intervals covered the final incidence in calendar week 8 for all six age groups, 50% prediction intervals for four out of the six age-groups (Fig. 2B).

Figure 3 shows results of the nowcasting for total incidence for six additional weeks during the ARE season 2024/2025. Nowcasting enabled an adequate qualitative real-time assessment of the current situation and trends during all phases of the epidemic, both before the wave (calendar weeks 45 and 50, 2024), during the sharp increase in SARI incidence (calendar week 4, 2025), after the peak of the wave (calendar week 10, 2025), and in the phase of declining incidence (calendar weeks 13 and 18, 2025).

**Figure 3.**
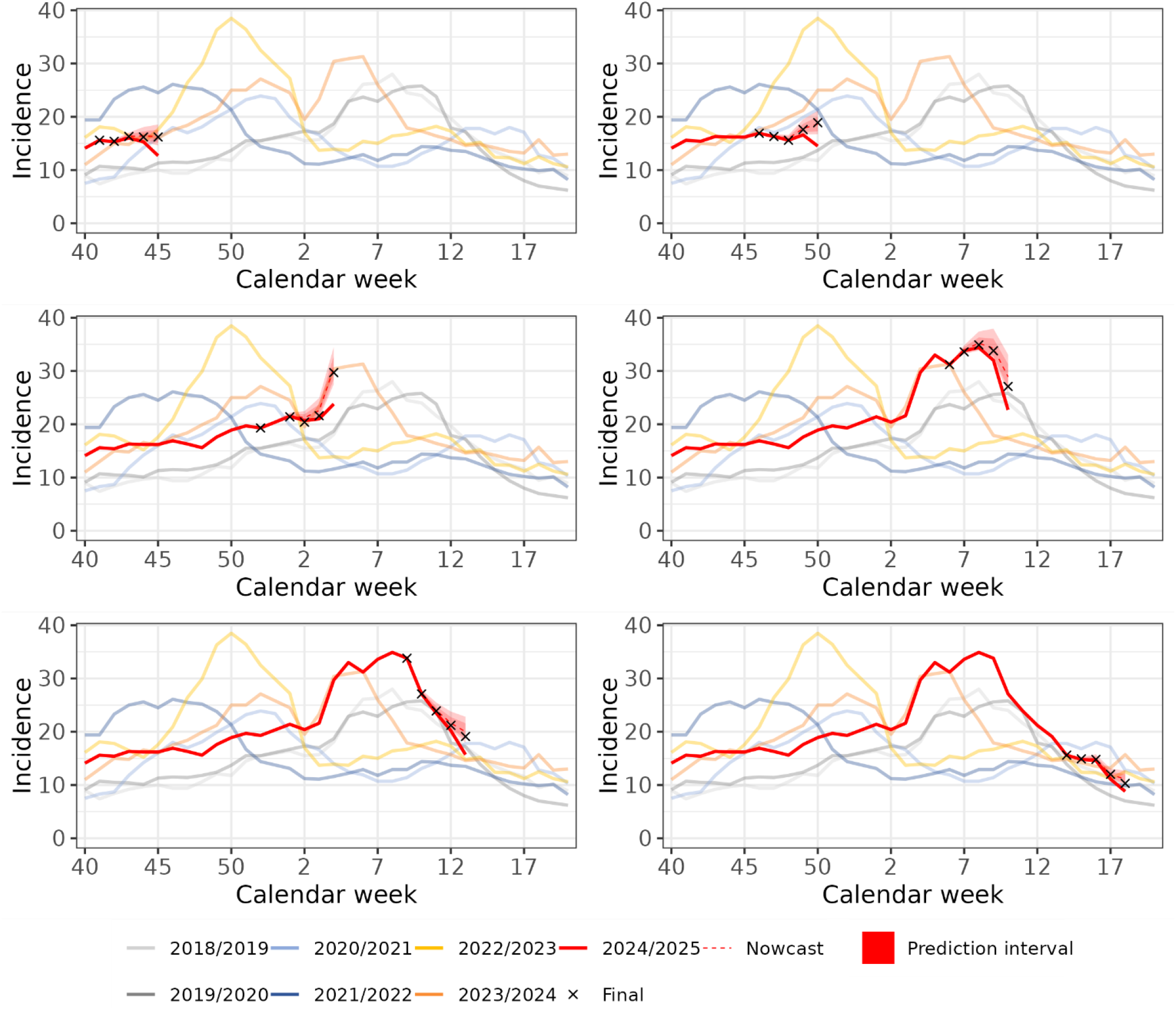
Illustration of the SARI nowcasting results for different calendar weeks and epidemic situations throughout the ARI season (total). Shown is the reported incidence data by calendar weeks in years up until 6 different weeks during the 2024/2025 ARI season representing different epidemic phases (incidence estimation up until calendar weeks 45 and 50, 2024, as well as calendar weeks 4, 10, 13, and 18, 2025, always based on data available in the subsequent week, i.e., the first week after hospitalisation). Also shown is the corresponding nowcast estimate of the expected final incidence (red dotted lines) and associated 80% and 95% prediction intervals (ribbons). Black crosses show the final incidence time series that was unavailable at the time of nowcasting.

### Performance evaluation of the nowcast

The proposed nowcasting method performed well over the respiratory infection season 2024/2025, with real-time predictions in the initial reporting week matching the final SARI incidence closely (Figure 4A). The nowcasts for the calendar weeks 2, 3, 5, and 6 in year 2025 overestimated the final SARI incidence slightly in the first week after hospitalisation (Figure 4A). Nevertheless, the nowcasted incidence trends improved qualitative assessment of current trends compared to the preliminary SARI hospitalisation incidence data available in those weeks (SuppFig. 1). Compared to the nowcast predictions in the first week after hospitalisation, prediction accuracy of the final incidence increased as more data accumulated (two, three, or four weeks after hospitalisation, SuppTab. 1). The model-based prediction intervals were well-calibrated, i.e., empirical coverage frequencies were close to or slightly above the desired levels (Figure 4B).

**Figure 4.**
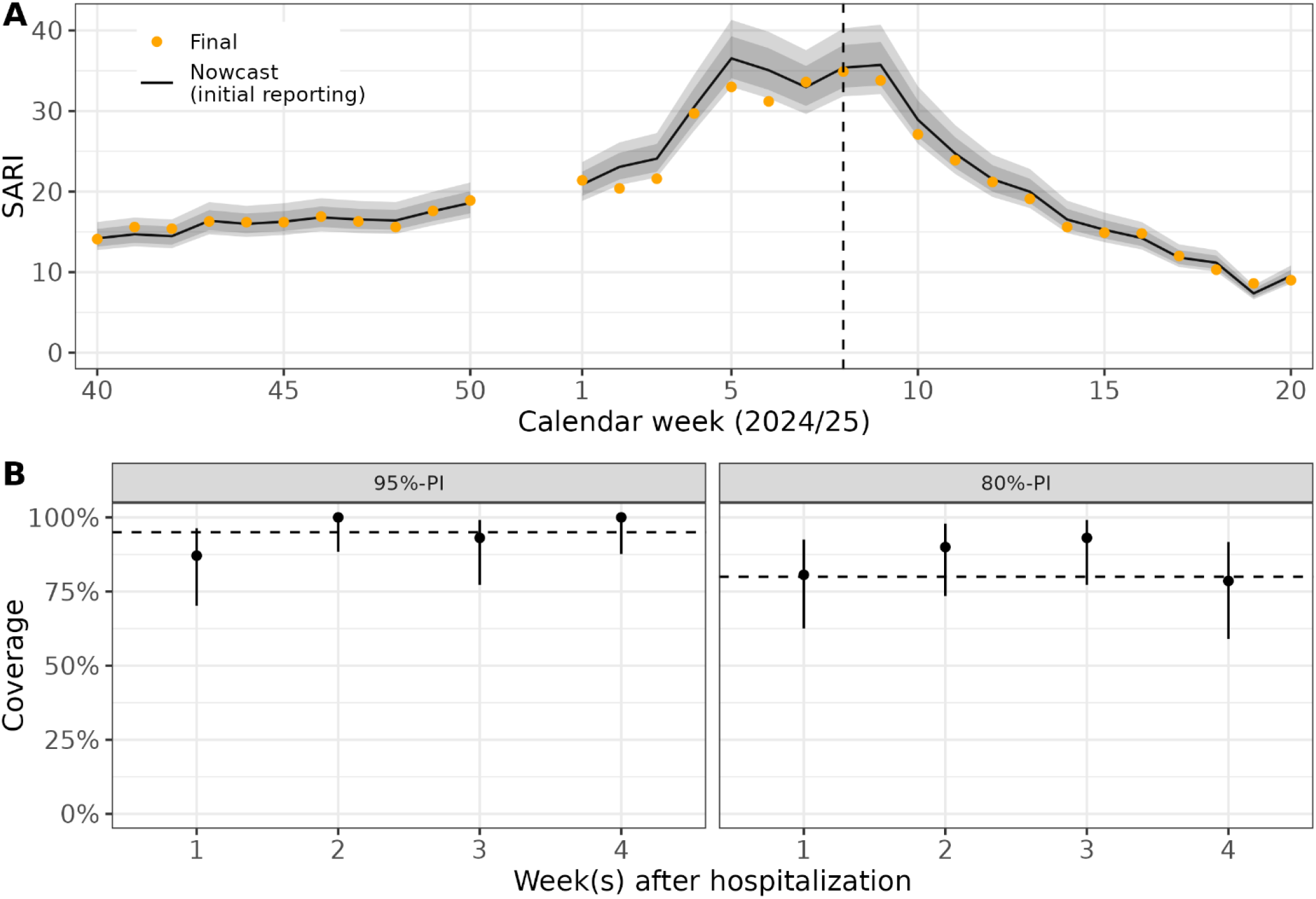
Retrospective evaluation of SARI nowcasting during the 2024/2025 ARE season. Panel A shows the reported nowcast prediction of the final SARI incidence for the week of initial reporting from applying the nowcast method to every week from calendar week 40, 2024 to calendar week 20, 2025 (black line) plus associated 80% and 95% prediction intervals (grey ribbons) based on data available in the subsequent week, i.e., the first week after hospitalisation. Orange dots show the actual final incidence time series. The black dotted lines indicate calendar week 8, 2025, previously used as example for illustrating the nowcasting method (cf. Figure 2). Panel B show the empirical coverage frequencies of 95% and 80% prediction intervals during the evaluation period for the nowcast 1-4 weeks after hospitalisation. Error bars correspond to associated 95% confidence intervals for the binomial proportion, dotted lines represent the desired coverage frequencies of the respective intervals.

We also performed the evaluation in each of the six age groups. We did not observe any problems when applying the nowcasting method to such smaller subgroups, albeit relative uncertainty around the nowcast point estimates increased for age groups with smaller incidence (e.g., age groups 15-34 years, and 35-59 years, SuppFig. 2). Relative errors in the point estimates (scaled mean absolute percentage error, sMAPE) were lowest in age groups with highest incidence (0-4 and 80+ years). Prediction intervals were well-calibrated in all age groups (cf. SuppTable 2; also, for additional performance measures).

## Discussion

In this work, we aimed to systematically investigate the magnitude of right-truncation in the SARI hospitalisation incidence and develop a robust nowcasting method to adjust for the truncation and the changes of the incidence data following their initial reporting. We evaluated the performance of the model during the 2024/2025 ARI winter season and investigated the benefits of applying the methods for real-time monitoring. Mostly, truncation of the data lead to a downward bias in incidence estimates for the most recent weeks (one or two weeks after hospitalisation, as a certain fraction of incident cases get their SARI diagnosis some weeks after their hospitalisation. These retrospective changes can occur due to pending diagnoses or due to review of preliminary primary and secondary diagnoses upon discharge. Our investigation revealed, that changes in incidence estimates were relatively stable throughout the winter season 2024/2025. While around 80 % of incident cases were available already in the first week after hospitalisation and 95 % in the second, the surveillance system already allows for a relatively fast assessment of the current situation. This is made possible by the use of electronic health records ^16,19^, as well as largely automated and digital data transmission and processing, as well as the use of preliminary diagnosis codes. In other surveillance systems, that can not rely on electronic health records, changes can occur over substantially longer periods of time. Within the German notification surveillance system, several steps are needed within the public health authorities, until data is reported to RKI: from laboratory confirmation, to case and contact management within the local health authorities, to federal health authorities and finally to the national health authority, the RKI^20^. For example, the estimation of COVID-19 hospitalisation incidence based on data from the German notification system during the COVID-19 pandemic showed substantially longer delays. However, part of this markedly larger delay was due to the aggregation of notified hospitalised cases by the date of first report of a COVID-19 case within the notification system, rather than the date of hospital admission^10^.

We applied our newly developed nowcasting method to the SARI sentinel data during the respiratory infections season 2024/2025 and we found that the results improved real-time interpretation of the incidence data overall as well as in subgroups by age. Specifically, the assessment of trend was very much improved during the 2024/25 influenza season, where the extent of increase or stabilisation can be difficult to evaluate. Moreover, older age groups tended to have a higher proportion of cases that were not known at the initial reporting week, thus the timely trend assessment in these age groups could be improved markedly. A retrospective evaluation of the performance showed that predictions of the nowcasting method were robust and the uncertainty quantification was well calibrated. The developed nowcasting method is conceptually elegant and straightforward and the results are available within minutes (on a regular laptop), also when being applied to multiple strata. Application to alternative stratified versions of the data, e.g., to specific subdiagnosis such as SARI caused by specific pathogens, is straightforward to implement, and we anticipate the model to perform well as long as the incidence is sufficiently high for a stable estimation.

Challenges arise in weeks with unusual data patterns, in particular during the Christmas and New Year holidays, when data were not processed due to a break in ARI reporting. For these weeks, the reporting patterns differed substantially compared to regular weeks, and the proposed method (that assumes stable reporting patterns) could not be applied. This is a known issue for data-driven statistical models using the empirical delay distribution^21^.

Our nowcasting method for the SARI hospitalisation incidence builds on weekly published time series of the estimated incidence counts based on data collected from multiple hospitals that are publicly available. Other data streams on disease incidence are available in similar data formats, and our proposed method is flexible enough to be applied to these as well. For the ICD-10 code based SARI hospital sentinel, a possible alternative would be to adjust the hospital-specific case counts directly for truncation and to perform the SARI incidence estimation based on these adjusted case counts. This approach was not part of the present work, and future research may examine whether this would improve the estimation accuracy. However, the hospital-level data is subject to strict data protection requirements and data sharing agreements.

In comparison to other European countries that have implemented a SARI surveillance system, the German SARI surveillance is within the range of other systems, that have the majority of the cases available within the first two weeks of reporting. However, the time to the final completion of clinical data entry is mostly much longer than the first availability of preliminary data also in other countries. In the Norwegian SARI surveillance system, nowcasting was also used to improve timeliness further^16,22-25^.

## Conclusion

The developed nowcasting method was shown to yield robust and reliable results. Hence, it seems justified to assume that the approach will work for different surveillance systems too, which we will explore next. The work also benefitted from the collaborative exchange as part of the RESPINOW project, which developed further ideas for collaborative real-time modelling of respiratory diseases in Germany^21^. Our experience from applying the method during the previous ARI season in winter 2024/2025 showed that the nowcasting can increase interpretability of the SARI hospitalisation incidence data substantially and we thus plan to incorporate the nowcasting into routine reporting of the indicator.

## Data Availability

The weekly SARI incidence estimate from the hospital sentinel is published year-round in a public repository (doi:https://doi.org/10.5281/zenodo.8382330).

## Acknowledgments

We thank our cooporation partner the HELIOS Kliniken GmbH and all sentinel hospitals for participating in the sentinel as well as work on data extraction, data transmission and data base programming, specifically Florian Hammerschmidt. We would like to thank Daniel Wesseler from Robert Koch Institute for technical support as well as maintenance and further development of the database.

