## Supplemental Material for "Development and implementation of a nowcasting method for the syndromic surveillance of severe acute respiratory infections (SARI) in Germany"

**Title**

### Supplementary material

**Supplementary Text 1: Parametric mixture distribution for modelling multiplicative scaling factors**

To model changes in the estimated incidence reported with a fixed time-lag of $d = 1, \dots, D-1$ weeks after hospitalization, we estimate a parametric distribution to J observed multiplicative changes of incidence, $\kappa_{t,d} = I_{t,d}/I_{t,D}$, observed in the past. The type of distribution we fit to the multiplicative factors depend on the available data:

1. If all $\kappa_{t,d}$ lie between (i.e., in the open interval) $(0, 1)$, we estimate parameters of a standard Beta(p, q) distribution via Maximum likelihood based on the observed J data points $\kappa_{t,d}$. For sampling of the $l = 1, \dots, L$ scaling factors, we sample first a realization of the parameter vector $\theta_d^{l,*} = (p_d^{l,*}, q_d^{l,*})$ from the asymptotic multivariate normal distribution of the maximum likelihood estimator, and draw then a random $k_{l,d}^*$ from the corresponding $Beta(p_d^{l,*}, q_d^{l,*})$ distribution.
2. If some of the observed $\kappa_{t,d}$ equal one and all other lie between $(0, 1)$, we use a one-inflated beta distribution and proceed otherwise as described above. The parameter vector $\theta$ contains in this case an additional parameter representing the fraction of $\kappa_{t,d}$ being equal to one.
3. If some of the observed $\kappa_{t,d}$ are bigger than one, we split the observed data into two parts: All $\kappa_{t,d} \leq 1$, and the $\kappa_{t,d} > 1$. For the former, we proceed as described in 1.) and 2.). For the $\kappa_{t,d} > 1$, we estimate a Beta distribution to the inverse $1/\kappa_{t,d}$, in the same way as described in 1.). To sample the $l = 1, \dots, L$ scaling factors for nowcasting, we first draw a random realization of the (binomial) probability of scaling factors being larger than one $\hat{\pi}_1$, and then draw $\hat{\pi}_1 \times L$ samples from the fitted Beta distribution of the inverse multiplicative changes (based on the $\kappa_{t,d} > 1)$ $and$ $(1 - \hat{\pi}_1) \times L$ samples from the fitted (one-inflated) Beta distribution (based on all $\kappa_{t,d} \leq 1)$, as described above.

**Supplementary Figure 1: Detailed visualization of selected nowcasts from the evaluation period.**

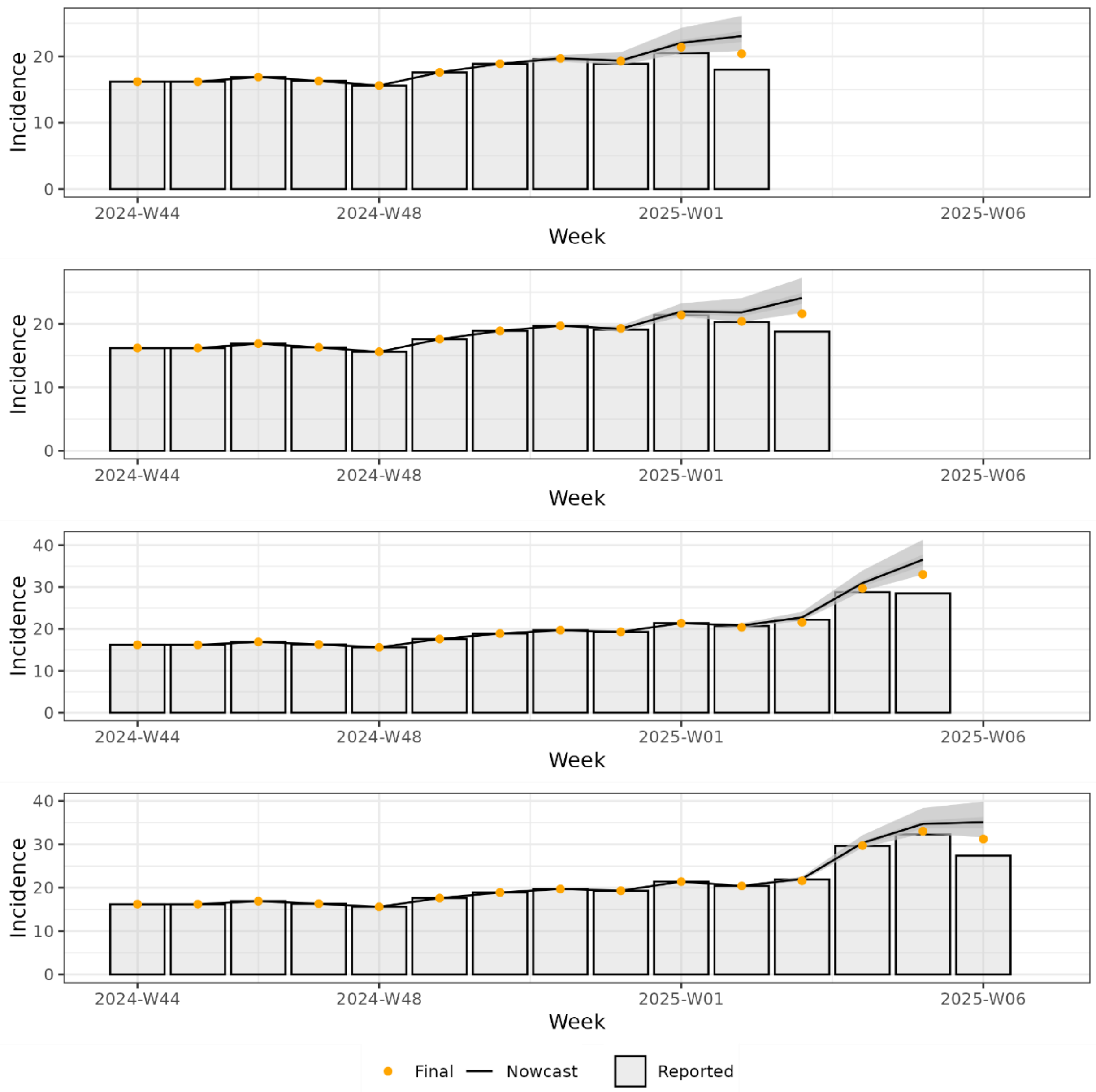

**Supplementary Table 1: Results of the retrospective evaluation of the nowcasting method for the overall SARI incidence in Germany during the winter season 2024/2025 (calendar week 40, 2024 until calendar week 20, 2025).**

| Week after hospitalization | MAE | RMSE | sMAPE | Mean WIS | Coverage PIs | | |
|---|---|---|---|---|---|---|---|
| | | | | | 95% | 80% | 50% |
| 1 (initial reporting) | 0.94 | 1.36 | 4.58 | 0.48 | 0.87 (0.7-0.96) | 0.81 (0.63-0.93) | 0.52 (0.33-0.7) |
| 2 | 0.48 | 0.64 | 2.18 | 0.21 | 1 (0.88-1) | 0.9 (0.73-0.98) | 0.73 (0.54-0.88) |
| 3 | 0.23 | 0.36 | 1.08 | 0.14 | 0.93 (0.77-0.99) | 0.93 (0.77-0.99) | 0.83 (0.64-0.94) |
| 4 | 0.15 | 0.2 | 0.7 | 0.08 | 1 (0.88-1) | 0.79 (0.59-0.92) | 0.61 (0.41-0.78) |

MAE: mean absolute error, RMSE: root mean squared error, sMAPE: scaled mean absolute percentage error, WIS: weighted interval score, Coverage PIs: empirical share of prediction intervals that contained final incidence and associated binomial 95% confidence intervals.

**Supplementary Figure 2: Evaluation of nowcast performance during respiratory infection season 2024/2025 in age groups.**

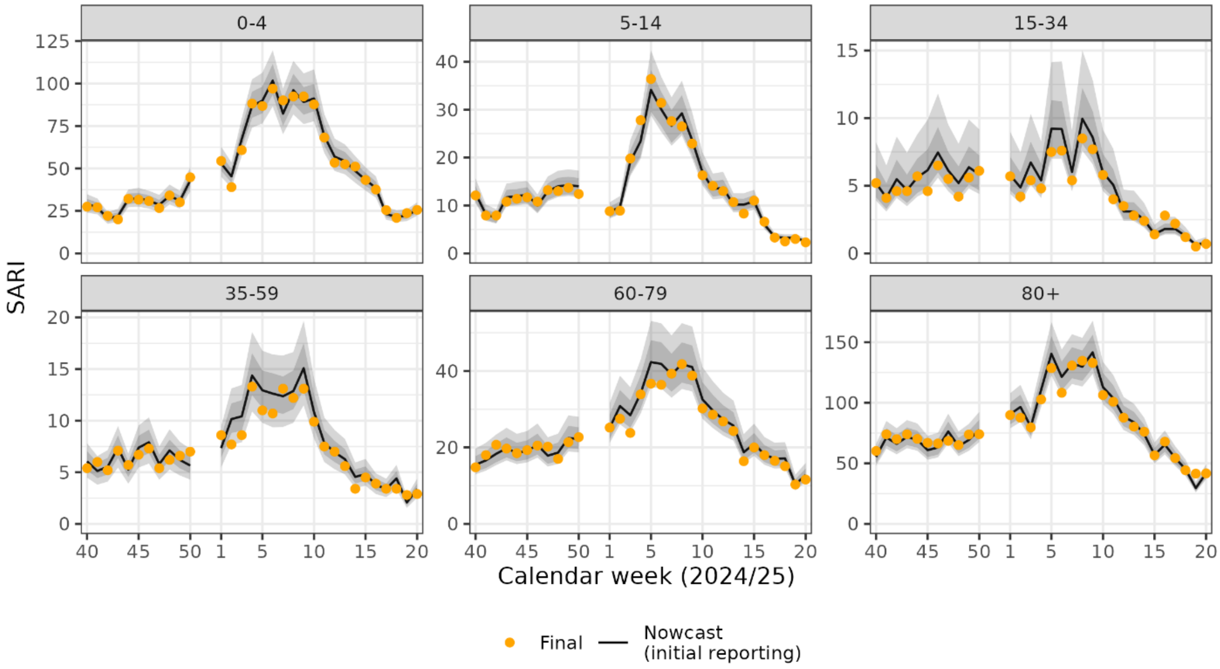


Shown are nowcast predictions (black lines) and associated 50% and 95% prediction intervals (grey ribbons) in the weeks of initial reporting. Orange dots correspond to the final incidence in each week. Note the different y-axis range for the different age-groups.

**Supplementary Table 2: Results of the retrospective evaluation of the nowcasting method for the SARI incidence in Germany during the winter season 2024/2025 (calendar week 40, 2024 until calendar week 20, 2025) in age groups.**

| Age group | MAE | RMSE | sMAPE | Mean WIS | Coverage PIs | | |
|---|---|---|---|---|---|---|---|
| | | | | | 95% | 80% | 50% |
| 00+ | 0.72 | 1.07 | 3.4 | 0.35 | 0.93 (0.84-0.98) | 0.85 (0.74-0.93) | 0.62 (0.49-0.74) |
| 0-4 | 1.57 | 2.3 | 3.31 | 0.82 | 0.98 (0.91-1) | 0.89 (0.78-0.95) | 0.66 (0.52-0.77) |
| 5-14 | 0.72 | 1.19 | 5.35 | 0.38 | 0.93 (0.84-0.98) | 0.87 (0.76-0.94) | 0.61 (0.47-0.73) |
| 15-34 | 0.43 | 0.61 | 9.45 | 0.19 | 0.97 (0.89-1) | 0.79 (0.66-0.88) | 0.61 (0.47-0.73) |
| 35-59 | 0.64 | 0.83 | 8.99 | 0.28 | 0.95 (0.86-0.99) | 0.7 (0.57-0.81) | 0.48 (0.35-0.61) |
| 60-79 | 1.27 | 1.81 | 5.07 | 0.61 | 0.98 (0.91-1) | 0.82 (0.7-0.91) | 0.59 (0.46-0.71) |
| 80+ | 3.35 | 4.51 | 4.19 | 1.64 | 0.98 (0.91-1) | 0.92 (0.82-0.97) | 0.62 (0.49-0.74) |

MAE: mean absolute error, RMSE: root mean squared error, sMAPE: scaled mean absolute percentage error, WIS: weighted interval score, Coverage PIs: empirical share of prediction intervals that contained final incidence and associated binomial 95% confidence intervals.

Results are aggregated over the nowcast predictions one and two weeks after hospitalization. The most robust results for comparing performance between age-groups is the relative error measure sMAPE, as absolute incidence levels vary between age-groups (see SuppFig. 2).
